# Rapid response to a measles outbreak in Ifanadiana District, Madagascar

**DOI:** 10.1101/2020.11.30.20143768

**Authors:** Karen E. Finnegan, Justin Haruna, Laura F. Cordier, Benedicte Razafinjato, Luc Rakotonirina, Andriamihaja Randrianambinina, Emmanuel Rakotozafy, Benjamin Andriamihaja, Andres Garchitorena, Matthew H. Bonds, Mohammed Ali Ouenzar

## Abstract

In 2019, Madagascar experienced the nation’s largest documented measles outbreak ever. From September 2018 to January 2020, nearly 225,000 individuals were infected, slightly more than 1,000 of whom died. Madagascar is one of the poorest countries in the world with one of the least funded health systems. Here, we present the experience of how Madagascar’s Ministry of Health (MoH) partnered with a non-governmental organization, PIVOT, to rapidly respond to a measles outbreak in the rural district of Ifanadiana, in the southeast of the country. The epidemic reached Ifanadiana in January of 2019. By August, there were more than 4,800 identified cases of suspected measles, including 157 hospitalizations. Over the course of two weeks in February 2019, the MoH and PIVOT mobilized nine teams of 220 total staff to vaccinate 69,949 children aged 6 months to 10 years at schools and community events. Surrounding the campaign, health workers were trained in measles identification and provided with medication for symptom management. The measles response required overcoming health system weaknesses and geographic barriers which are endemic throughout the country and create chronic challenges for the provision of routine preventive and curative care. This response demonstrates that rapid mobilization of an organized health system response is feasible even in hard-to-reach areas.

**KEY POINTS:**

- Madagascar experienced the island’s worst measles outbreak in history as a result of low vaccination coverage.
- In Ifanadiana District, the Ministry of Health and a non-governmental health organization collaborated to respond to the measles outbreak with a mass vaccination campaign, sensitization, and provision of supplies to health facilities for the management of disease complications.
- The measles outbreak in Madagascar highlights the need for an increased focus on routine preventive care, especially in hard-to-reach communities; calls attention to the consequences of ruptures in the care continuum; and highlights the need for timely, continuous, de-centralized epidemiologic data.
- The response to the outbreak in one rural district is evidence that health care can be delivered in remote areas.

## INTRODUCTION

Measles is a highly transmissible infectious disease which caused an estimated 110,000 deaths globally in 2017, the majority of which occurred in children under five.^1^ From September 2018 to January 2020, Madagascar grappled with the world’s worst measles epidemic in decades. Over 244,500 individuals were diagnosed, and 1080 died.

With no curative treatment available, the primary strategy for managing measles is through vaccination. Herd immunity can be reached if vaccination coverage rates exceed 95%.^2^ Despite longstanding and freely available national vaccination programs throughout the world, and increases in vaccination rates in the last two decades, coverage rates remain below critical thresholds for elimination in many priority countries.^2^ In Madagascar, measles vaccination coverage was estimated to be 58% in 2017, well below the 70% coverage estimated for sub-Saharan Africa and 95% required for herd immunity.^3,4^

The national vaccination protocol in Madagascar recommends that children receive the first dose of measles vaccine at nine months. Measles vaccines are provided to the Madagascar Ministry of Health (MoH) through partnership with multilateral agencies. Vaccines are distributed via bi-annual mass immunization campaigns and through routine immunization by health providers at primary care centers in every commune (administrative units which include 5,000-15,000 people) during designated weekly vaccination days. Although immunizations are free, they are not accessible to many because of both supply side factors (reliable cold chain, availability of health workers and commodities) and demand side factors such as geographic barriers to care (distance and terrain) and seasonal challenges to accessing care.^5^

In this report, we describe the 2018/2019 measles outbreak and response in the rural district of Ifanadiana, which is demographically similar to many other districts throughout the country.

## MEASLES OUTBREAK IN MADAGASCAR

Madagascar’s measles outbreak began in September of 2018. From September through the end of January 2020, there were 244,664 reported cases of measles. Nationally, 1,080 deaths were attributed to measles. The World Health Organization’s provisional estimate of measles incidence in Madagascar for the twelve month period prior to January 2020 was 5654.1 cases per 1,000,000 people, the highest in the world among countries reporting surveillance data for the same time period.^6^

### Setting

Ifanadiana District is located in the southeast region of Vatovavy-Fitovinany with an estimated population of 209,000 residing in 15 communes which comprise 195 fokontany (the smallest administrative unit). Each commune has at least one primary care health center which offers weekly vaccination services. A district hospital located in Ifanadiana commune provides advanced and emergency care. Ifanadiana District is bisected by a paved road, but 71% of the population resides more than five kilometers from a health facility and access to remote villages is limited during the rainy season. Most communities are accessible only by footpath through the hilly terrain.

PIVOT, an international non-governmental organization (NGO), began working in Ifanadiana District in 2014 to establish a district level model health system with the Ministry of Health. Its strategy for health system strengthening is through integrating clinical programs, facility readiness, and data systems at all levels of the health system in the district.^7–9^ PIVOT provides a basic support package to all health centers in the district, including funding to employ sufficient staff to meet MoH norms and joint supervision of facility-based staff collecting data as part of country’s Health Management Information System. In six health centers in the PIVOT catchment area, this basic support package is supplemented with improved infrastructure, payment of user fees through reimbursement of facilities, supply chain management, and support for the training of clinical staff. Additionally, PIVOT provides supervision for community health workers in the associated fokantany in coordination with the MoH. Ifanadiana District’s health system also receives more limited support from international aid agencies and NGOs in the form of funding, programming, consumables, and technical assistance.

The first case of measles in Ifanadiana District was identified in January 2019. During the height of the epidemic, the district health office shared case data with PIVOT to help coordinate the response between the MoH and NGO. From January to the end of July 2019, 4,819 cases of measles were reported with 157 hospitalizations and 38 reported deaths. Nearly 10% of measles cases occurred in children under one (Table 1). For the vast majority of cases (98%), measles vaccination status could not be confirmed upon diagnosis. Measles cases were identified at the health center or hospital and samples from suspected cases were sent to the national reference laboratory in Antananarivo for confirmation at the start of the epidemic. All health centers within the district and the hospital were required to submit weekly epidemiological surveillance data on number of suspected measles cases, number of confirmed cases, and the number of deaths due to possible measles at the health center and within the community.

**Table 1.** Characteristics of measles cases in Ifanadiana District

|  | N | % |
| --- | --- | --- |
| Male | 2395 | 49.7 |
| Female | 2426 | 50.3 |
| <1 year | 453 | 9.4 |
| Mean age in years (among those >=1 year) | 15.5 |  |
| Hospitalized | 157 | 3.3 |
| Mean time hospitalized (in days) among those hospitalized | 3.0 |  |

In 2018, prior to the start of the outbreak, measles vaccination coverage was 58% in Ifanadiana District (73% in the PIVOT catchment area and 49% in the rest of the district, p<0.01), based on data from a representative longitudinal cohort selected in 2014.^10^ The district coverage estimate was consistent with national coverage.

### Response to the outbreak

Nationally, the MoH convened leadership and partner organizations to identify available resources and plan a response to the outbreak. Vaccination campaigns were scheduled for a staged roll out throughout the country due to limited availability of funding and personnel. With additional logistical and financial support, the MoH and PIVOT launched an expedited campaign in the third week of February 2019. Prior to the campaign launch, there were widespread sensitization activities on the importance of vaccination and locations to receive the services. Before commencing vaccination activities, PIVOT ensured that all health facilities in the district were equipped with medication for treatment of measles symptoms and that clinicians were trained on signs and symptoms, diagnosis and treatment.

Vaccines and staff were transported from Antananarivo by PIVOT vehicles and the vaccines were stored before being dispatched in cold boxes in vehicles or on motorcycles. Vaccination activities were conducted by nine teams, which included the head of the commune’s health center, a representative from the MOH district health office, and a PIVOT staff member. In total, 220 vaccinators were involved in addition to 110 community health workers. Due to impassable terrain, more than 50% of people vaccinated were reached by teams traveling by foot; others were reached via motorcycle.

In total 74,589 vaccine doses were transported from Antananarivo to Ifanadiana District. Vaccination events targeted children aged 6 months to 10 years and were held in primary schools, community health posts, and at community events. Over five days, 61,208 children were vaccinated with an additional 8,741 vaccinated during follow-up events for a total of 69,949 children vaccinated. Any child with a fever was referred to the nearest health facility for diagnosis and received a vaccination following 48 hours fever-free. Health facilities proactively reviewed vaccination cards of all children presenting for care and encouraged vaccination of children who were not fully vaccinated against measles.

Following the campaign, PIVOT supported MoH clinicians in the diagnosis and treatment of measles cases at health facilities, provided an ambulance for emergency transfers from the health center to the hospital, and supported the reporting of surveillance data from facilities to the district office. These are routine activities of PIVOT.

Ifanadiana District was declared free of measles cases in November 2019.

## REFLECTION

The measles outbreak in Madagascar highlights the importance of continued focus on preventive care—specifically vaccination—in improving population health. Funding of siloed vertical disease programs, even those which focus on the health of vulnerable populations, have been inadequate in preventing disease outbreaks. With a strengthened health system, and collaboration of partners, rapid responses are feasible.

The implementation of a responsive district-wide vaccination campaign in Madagascar is possible. In Ifanadiana District, vaccination efforts were constrained by geographic access and the condition of roads. Despite efforts to overcome geographic barriers, there were remote communities which were unable to be reached for vaccination events due to impassable paths or waterways. Additionally, measles vaccination activities had to account for health system gaps such as poor facility infrastructure, lack of running water and unreliable energy. These infrastructure challenges also detract from other clinical services offered at facilities, including treatment of sick children, antenatal care visits, and deliveries.

Although the MoH and PIVOT were able to mount a rapid response to the epidemic, vaccination efforts were not sustained with the same fervor following the first round of the vaccination campaign. As the immediate needs of the population were addressed through targeted mass vaccination, attention shifted to identification and treatment of cases. Prevention efforts should be sustained through ongoing, increased attention on routine vaccination programs.

Inadequate funding, a health system buffeted by recurrent plague, and persistent malnutrition among children under-five, strengthened the measles outbreak.^11–13^ PIVOT provides support to a subset of health facilities in Ifanadiana District with plans to expand to all facilities by 2022. Despite implementation of a health systems strengthening intervention which targets all levels of the health system (community, health center, and hospital), vaccination coverage remains below global targets; as noted previously, measles coverage was 73% in the PIVOT catchment area in 2018 and 49% in the rest of the district. The epidemic highlights the importance of universal coverage and consistent access to high quality health care. Although increases in vaccination coverage have been achieved in Ifanadiana and the rest of the country in a short time in response to the epidemic, the only way to prevent future outbreaks is to strengthen routine vaccination and preventive care over the long term.

The delivery of routine preventive and curative care is essential in serving the care continuum. Geographic barriers, lack of trust in the health system, inability to pay, and an unreliable health system, limit access.^5,7,14^ A resilient and responsive health system provides preventive and curative care which includes high quality antenatal care, facility-based delivery by a trained health provider, postnatal care, and on-time vaccination of children. These components of the care continuum require adequate staffing, supply chain, infrastructure, and medicines and commodities. A rupture in the system often gives rise to recurrent emergencies (e.g. plague in 2017, measles in 2018) which requires a rapid response that further diverts resources from the primary care system.

The measles outbreak also highlights the importance of continuous epidemiologic surveillance data for outbreak identification and response. The availability of timely and high quality data are essential for preparing an informed response to an epidemic, especially in contexts with limited resources.

## Conclusion

In collaboration with the Madagascar Ministry of Health, PIVOT oversaw the administration of nearly 70,000 doses of measles vaccine in Ifanadiana District over a five-day period. Health care delivery in the district, as in much of Madagascar must grapple with limited resources, a health system challenged by a high disease burden, and geographical barriers which restrict access to remote communities. However, the vaccination campaign in the district demonstrates that collaboration and pooling of district, donor, and non-governmental organization resources can result in rapid intervention coverage potentially sufficient for herd immunity even when confronted with an epidemic.

## Data Availability

The corresponding author can provide information on the process of accessing the data.

## ACKNOWLEDGMENTS

We would like to thank the frontline health workers, including doctors, nurses, and community health workers who responded to the measles epidemic. We thank the Ministry of Health staff, colleagues at World Health Organization, and other members of the national epidemic response team for their collaboration during the epidemic.

## COMPETING INTERESTS

KEF, JH, LFC, BR, LR, BA, MHB, and MAO received salary support from PIVOT.

## FUNDING

The authors have not declared a specific grant for this research from any funding agency in the public, commercial or not-for-profit sectors.

---

**Figure 1.**
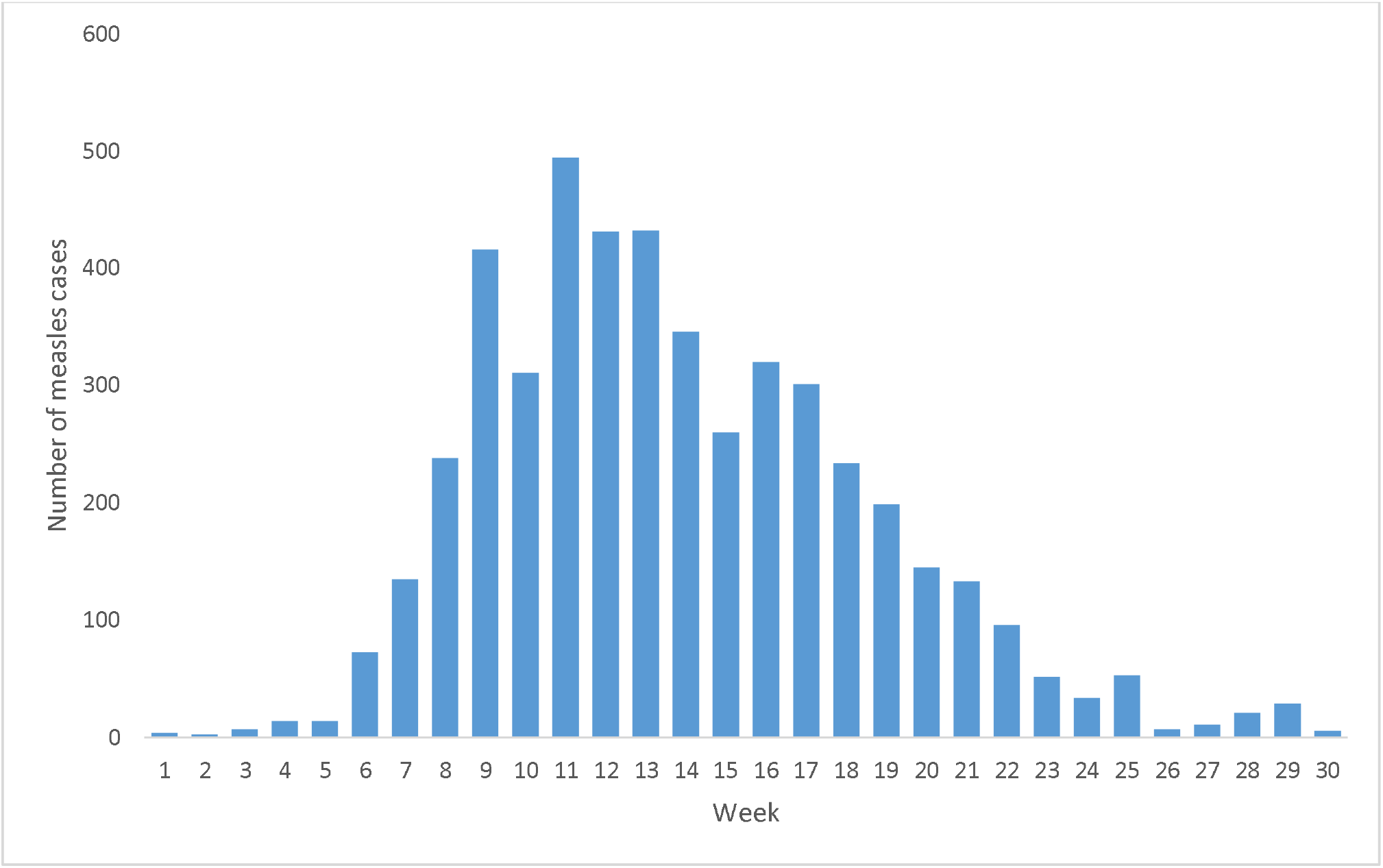
Epidemiologic curve of cases diagnosed by week in Ifanadiana District (January - July, 2019)

## Notes

### Competing Interest Statement

KEF, JH, LFC, BR, LR, BA, AG, MHB, and MAO received salary support from PIVOT. AR and ER received salary support from Madagascar's Ministry of Public Health.

### Funding Statement

No external funding was received for the preparation of this manuscript. Program activities which are reported on were funded by grants and donations received by PIVOT.

### Author Declarations

This work uses publicly available data and was authorized by the leadership of Madagascar's Ministry of Public Health. This study was determined to be non human subjects research by the IRB at Harvard University.

